# Length of hospital stay and risk of intensive care admission and in-hospital death among COVID-19 patients in Norway: a register-based cohort study comparing patients fully vaccinated with an mRNA vaccine to unvaccinated patients

**DOI:** 10.1101/2021.11.05.21265958

**Authors:** Robert Whittaker, Anja Bråthen Kristofferson, Beatriz Valcarcel Salamanca, Elina Seppälä, Karan Golestani, Reidar Kvåle, Sara Viksmoen Watle, Eirik Alnes Buanes

**Author notes:** Corresponding author: Robert Whittaker, Norwegian Institute of Public Health, Lovisenberggata 8, 0456, Oslo, Norway.

## Abstract

**Objectives:** With most of the Norwegian population vaccinated against COVID-19, an increasing number and proportion of COVID-19 related hospitalisations are occurring among vaccinated patients. We estimated the length of stay (LoS) in hospital and an intensive care unit (ICU), and risk of admission to ICU and in-hospital death among COVID-19 patients ≥18 years who had been fully vaccinated with an mRNA vaccine, compared to unvaccinated patients.

**Methods:** Using national registry data, we conducted a cohort study on SARS-CoV-2 positive patients hospitalised in Norway between 1 February and 30 November 2021, with COVID-19 as the main cause of hospitalisation. We ran Cox proportional hazards models to analyse differences in our outcomes. Explanatory variables included vaccination status, age, sex, county of residence, regional health authority, date of admission, country of birth, virus variant and underlying risk factors.

**Results:** We included 3,203 patients, of whom 716 (22%) were fully vaccinated (at least two doses or one dose and previous SARS-CoV-2 infection). Fully vaccinated patients had a shorter overall LoS in hospital (aHR for discharge: 1.61, 95%CI: 1.24–2.08), shorter LoS without ICU (aHR: 1.27, 95%CI: 1.07–1.52), and lower risk of ICU admission (aHR: 0.50, 95%CI: 0.37–0.69) compared to unvaccinated patients. We observed no difference in the LoS in ICU, nor risk of in-hospital death between fully vaccinated and unvaccinated patients.

**Conclusions:** Fully vaccinated patients hospitalised with COVID-19 in Norway have a shorter LoS and lower risk of ICU admission than unvaccinated patients. These findings can support patient management and ongoing capacity planning in hospitals.

## Introduction

COVID-19 vaccination programmes have drastically reduced the burden of COVID-19 related hospitalisations and deaths (1-5). However, the risk of breakthrough cases of severe COVID-19 after vaccination remains, particularly among groups at higher risk of severe disease (6, 7).

Norway (population 5.4 million) started COVID-19 vaccination in December 2020, initially focusing on individuals ≥65 years, health care workers and individuals at increased risk of severe COVID-19 (8). The mRNA vaccines Comirnaty® (BioNTech-Pfizer, Mainz, Germany/New York, United States) and Spikevax® (mRNA-1273, Moderna, Cambridge, United States) are the two predominant vaccines administered (9). National second dose coverage among ≥18-year-olds reached 87% by 30 November 2021. Persons with specific immunosuppressive conditions were offered a third dose as part of the primary series from September (10). Booster doses have been offered to persons ≥65 years and care home residents since October, and health care workers since November 2021 (11).

With high national vaccination coverage, an increasing number and proportion of COVID-19 related hospitalisations are occurring among vaccinated patients, characterised by advanced age and underlying comorbidities that increase the risk of severe COVID-19 (8, 12). It is therefore essential to understand how vaccination may affect clinical endpoints among patients who are hospitalised for COVID-19 to support patient management and capacity planning in hospitals. Published data on this are currently limited (13, 14).

We linked individual-level data from national registries to estimate the length of stay (LoS) in hospital (with and without intensive care unit (ICU) stay) and ICU, and risk of ICU admission and in-hospital death among COVID-19 patients ≥18 years in Norway who had been fully vaccinated with an mRNA vaccine, compared to unvaccinated patients.

## Methods

### Patient cohort

We conducted a cohort study on patients ≥18 years hospitalised between 1 February and 30 November 2021 after a positive SARS-CoV-2 test and who had a national identity number registered. We included patients hospitalised ≤2 days before and ≤28 days following a positive SARS-CoV-2 test, where COVID-19 was the reported main cause of hospitalisation. Cases hospitalised with other or unknown main cause were excluded. We did not restrict admissions by LoS. The Alpha variant was the predominant circulating variant at the start of the study period, before being superseded by Delta in early July (15).

### Data sources

We obtained data from the Norwegian national emergency preparedness registry for COVID-19 (16). This registry contains individual-level data on all laboratory-confirmed COVID-19 cases, COVID-19 related hospitalisations and ICU admissions, and COVID-19 vaccinations among Norwegian residents. Further details on the data sources and national COVID-19 vaccination coverage over time by age group and number of doses, are presented in supplementary materials A, part 1. We extracted data from the preparedness registry on 14 December 2021, ensuring a minimum of 13 days follow-up since last date of hospitalisation.

### Definition of COVID-19 vaccination status

Vaccination status was defined on the date of positive test for SARS-CoV-2:

1. Unvaccinated: Not vaccinated with a COVID-19 vaccine.
2. Fully vaccinated: Positive test ≥7 days after second dose with at least the absolute minimum interval between doses depending on vaccine type (17), or ≥7 days after first dose if previously diagnosed with a SARS-CoV-2 infection ≥21 days before vaccination.

We excluded patients vaccinated with only one dose, those who had received a second dose <7 days before date of positive test, and patients vaccinated with a non-mRNA vaccine only. We also excluded unvaccinated patients with reported reinfections of SARS-CoV-2.

### Outcome measures

Our outcomes were discharge from hospital (with and without ICU admission), admission to ICU, discharge from ICU and in-hospital death. We calculated LoS as the time between first admission and last discharge. For patients with more than one registered hospital stay, we included time between stays, if the time between two consecutive stays was <24 hours. For LoS in ICU, we included time between consecutive stays if <12 hours. Separate stays were registered if a patient was discharged and readmitted, or if a patient was transferred between wards or hospitals. Patients with unknown date of discharge from their last stay were considered to still be hospitalised. In-hospital death was registered at discharge.

### Data analysis

Explanatory variables used to analyse differences in our outcomes were vaccination status, age, sex, county of residence, regional health authority, date of admission, country of birth, virus variant and underlying risk factors (Table 1).

**Table 1.**
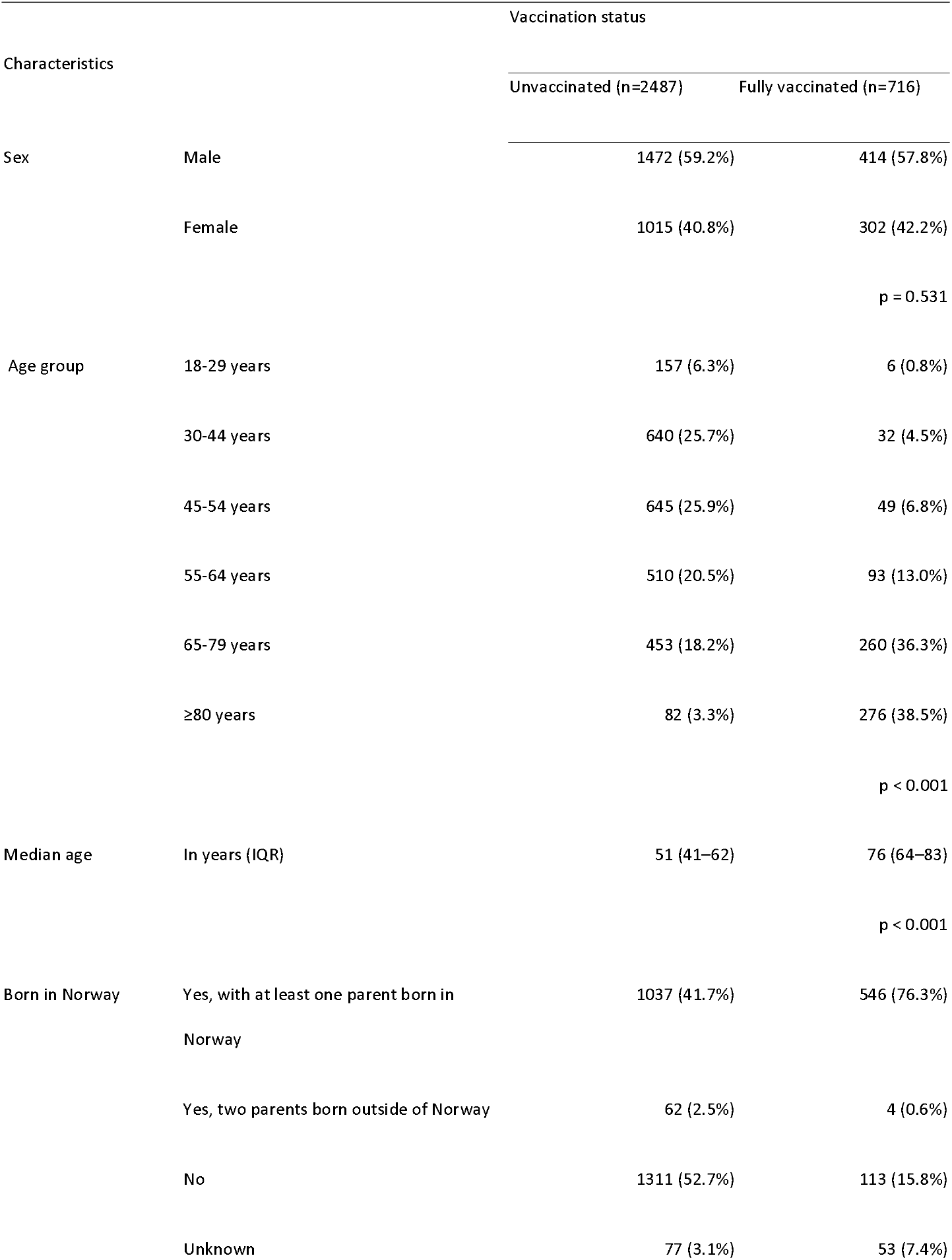
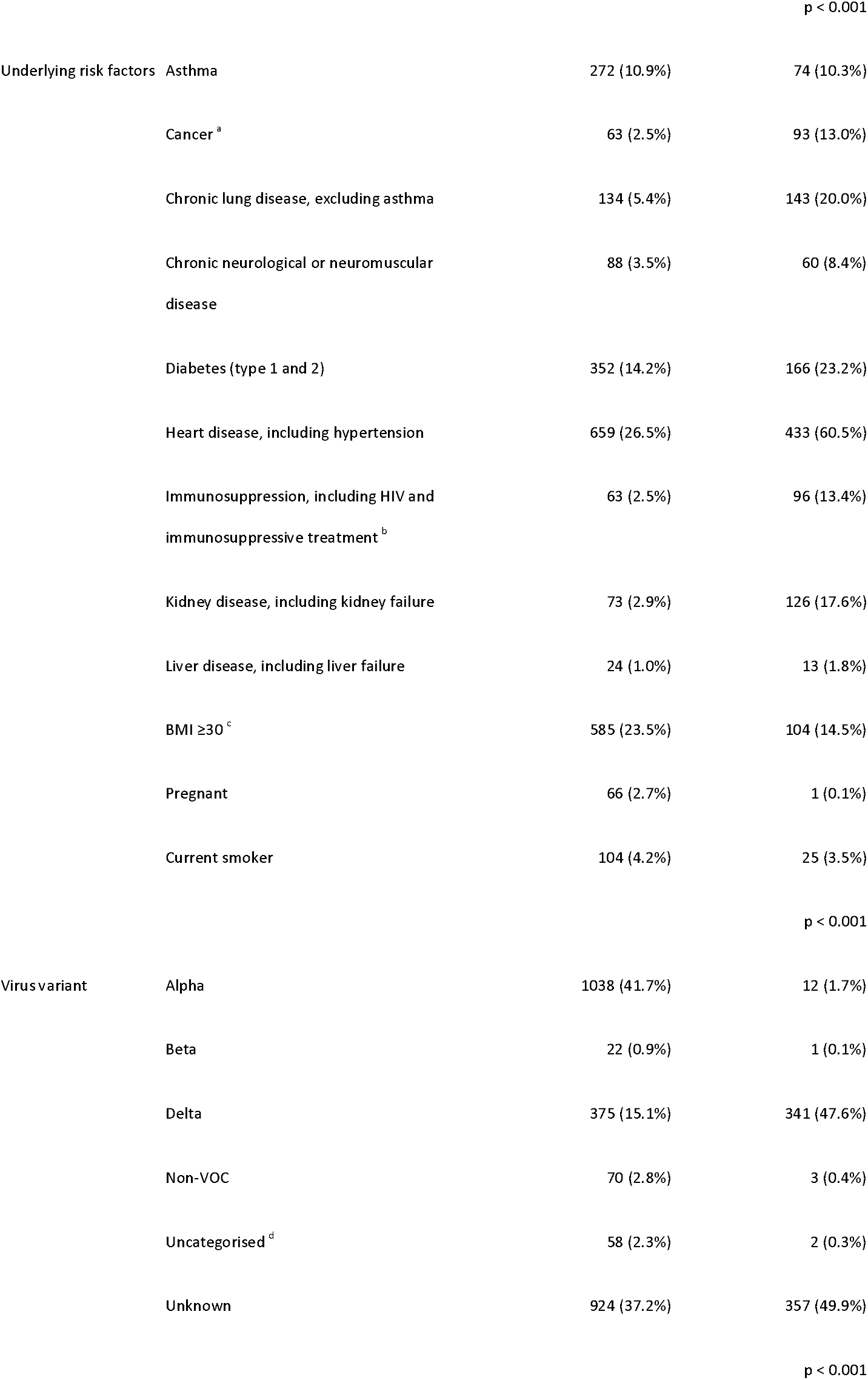
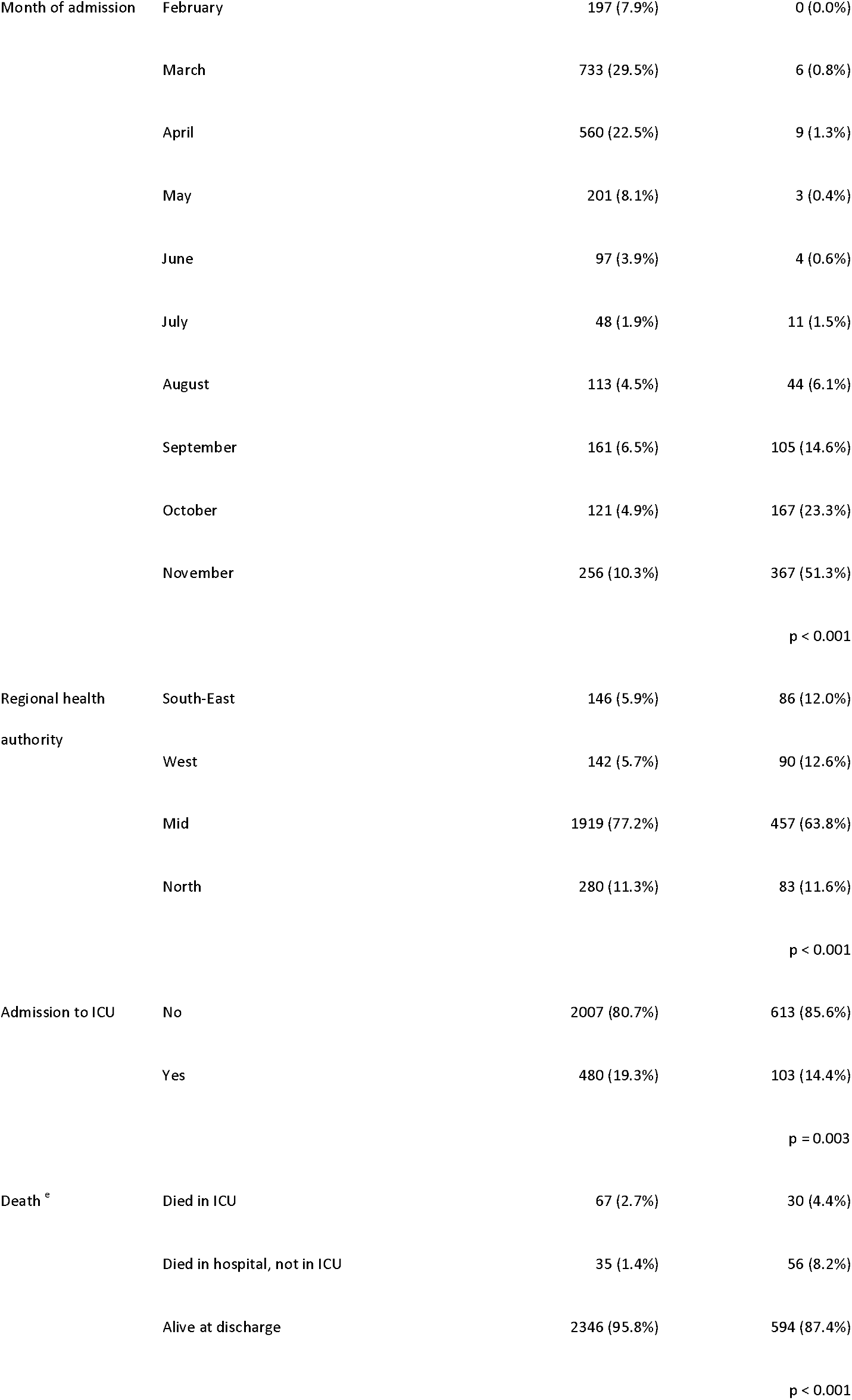
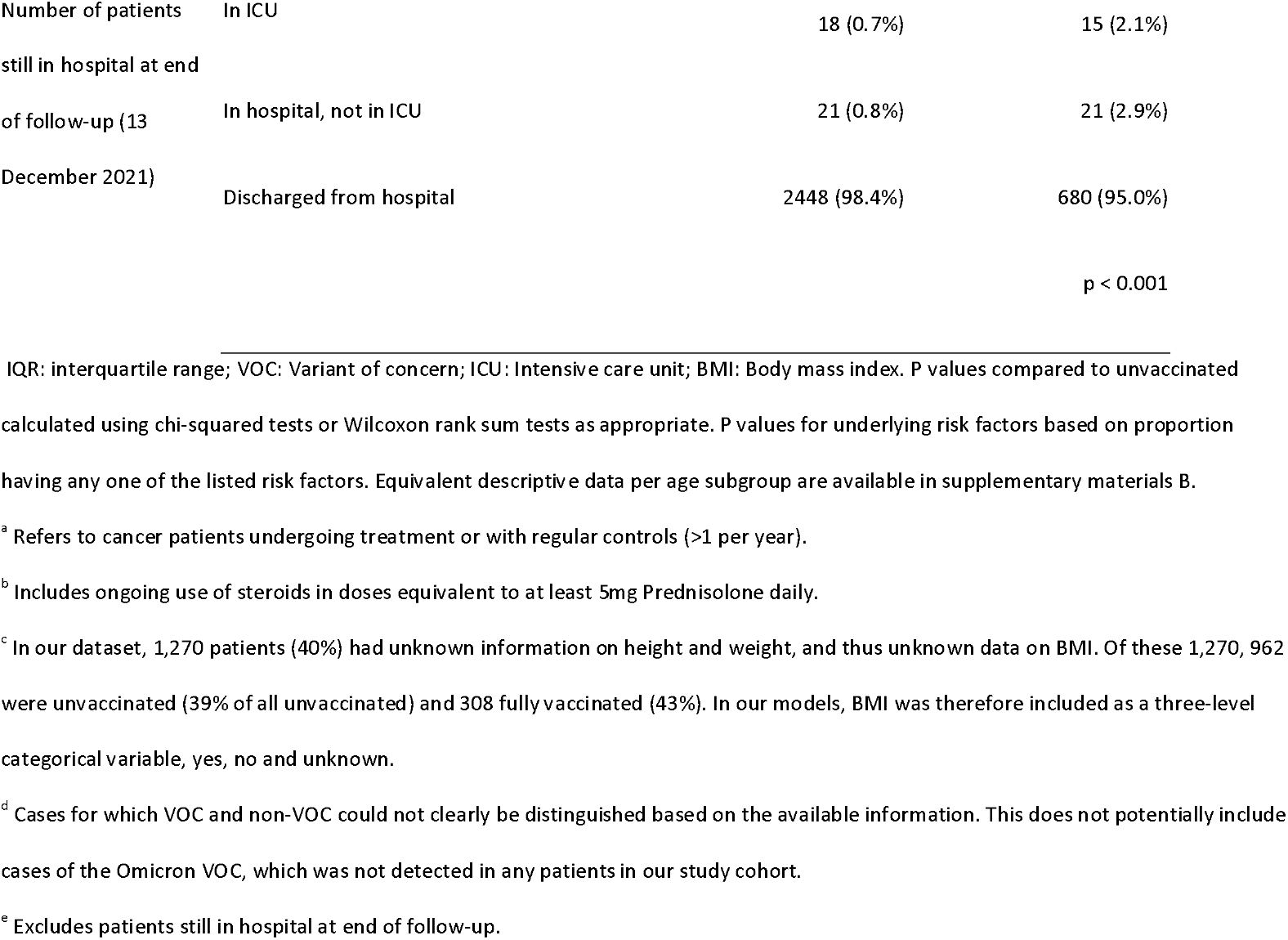
Characteristics of SARS-CoV-2 positive patients aged ≥18 years hospitalised with COVID-19 as the main cause of hospitalisation, by vaccination status, Norway, 1 February – 30 November 2021

Outcomes were first explored univariably in a Cox proportional hazards model and by calculating Kaplan Meier curves, with right censoring of patients still admitted to hospital. Crude log hazard ratios together with medians and interquartile ranges (IQR) for LoS were obtained. Explanatory variables with a p value <0.2 were further explored in multivariable models. Forward model selection was performed based on Akaike Information Criterion. Vaccination status was maintained in all models regardless of significance. Continuous variables (date of admission and age) were tested as linear, spline or categorically. The multivariable model was checked for the assumption of proportional hazard by checking Schoenfeld residuals, and some explanatory variables were stratified to satisfy the assumption. The adjusted log hazard ratios (aHR) obtained in the multivariable models were reported. We ran models on all patients ≥18 years, as well as for the following age subgroups: 18–64 years, 65–79 years and ≥80 years. Patients vaccinated with three doses were not analysed separately due to small numbers. LoS in ICU was not analysed for the age subgroups due to the small number of vaccinated patients admitted to ICU in each subgroup (≤50).

We also conducted sensitivity analyses by changing the definition of our study population, study period or our outcome definitions to further explore if our main results were robust (supplementary materials A, part 2). The statistical analysis was performed in R version 3.6.2.

### Ethics

Ethical approval for this study was granted by Regional Committees for Medical Research Ethics - South East Norway, reference number 249509. The need for informed consent was waived.

## Results

### Description of cohort

During the study period, 3,541 reported cases of COVID-19 were hospitalised with COVID-19 as the main cause of hospitalisation ≤2 days before and ≤28 days after a positive SARS-CoV-2 test. Of these, 3,476 (98%) had a national identity number registered. We excluded 262 patients vaccinated with only one dose or a second dose <7 days before date of positive test, four patients vaccinated with non-mRNA vaccines, one patient with unknown vaccine type and two unvaccinated patients who were reported as having been reinfected with SARS-CoV-2. We also dropped four patients who had a reported stay in ICU outside of their hospital stay, due to assumed incomplete reporting on hospital stays. Our study cohort included the remaining 3,203 patients.

The median time from positive test to hospitalisation was 5 days (IQR 1–8), and 3,157 (99%) patients were admitted within 14 days of positive test. In total, 583 (18%) patients were admitted to ICU. At the end of follow-up, 75 (2.3%) patients were still hospitalised. Of the 3,128 patients who had been discharged, 188 (6.0%) died in hospital. In total, 716 (22%) patients were fully vaccinated, of whom 666 (93%) had received two doses, 47 (6.6%) three doses and three (0.4%) one dose with a previous SARS-CoV-2 infection. Most patients (658, 92%) received a homologous Comirnaty regimen. A full breakdown of vaccine types and time between doses is presented in supplementary materials A, part 3. The median time from last dose to diagnosis was 174 days (IQR: 126–217). Age and the frequency of certain underlying risk factors such as cancer, chronic lung disease, heart disease, immunosuppression (due to illness or treatment) and kidney disease were higher among fully vaccinated patients. Detailed characteristics of the study cohort by vaccination status are presented in Table 1. Equivalent descriptive data per age subgroup are available in supplementary materials B.

### Length of stay in hospital and intensive care, and risk of admission to intensive care and in-hospital death by vaccination status

Descriptive data and crude and adjusted hazard ratios for each outcome by age subgroup and vaccination status are presented in Tables 2 and 3, and Figure 1. Estimates from all univariable and multivariable models in the main analysis are presented in supplementary materials B and C.

**Table 2.**
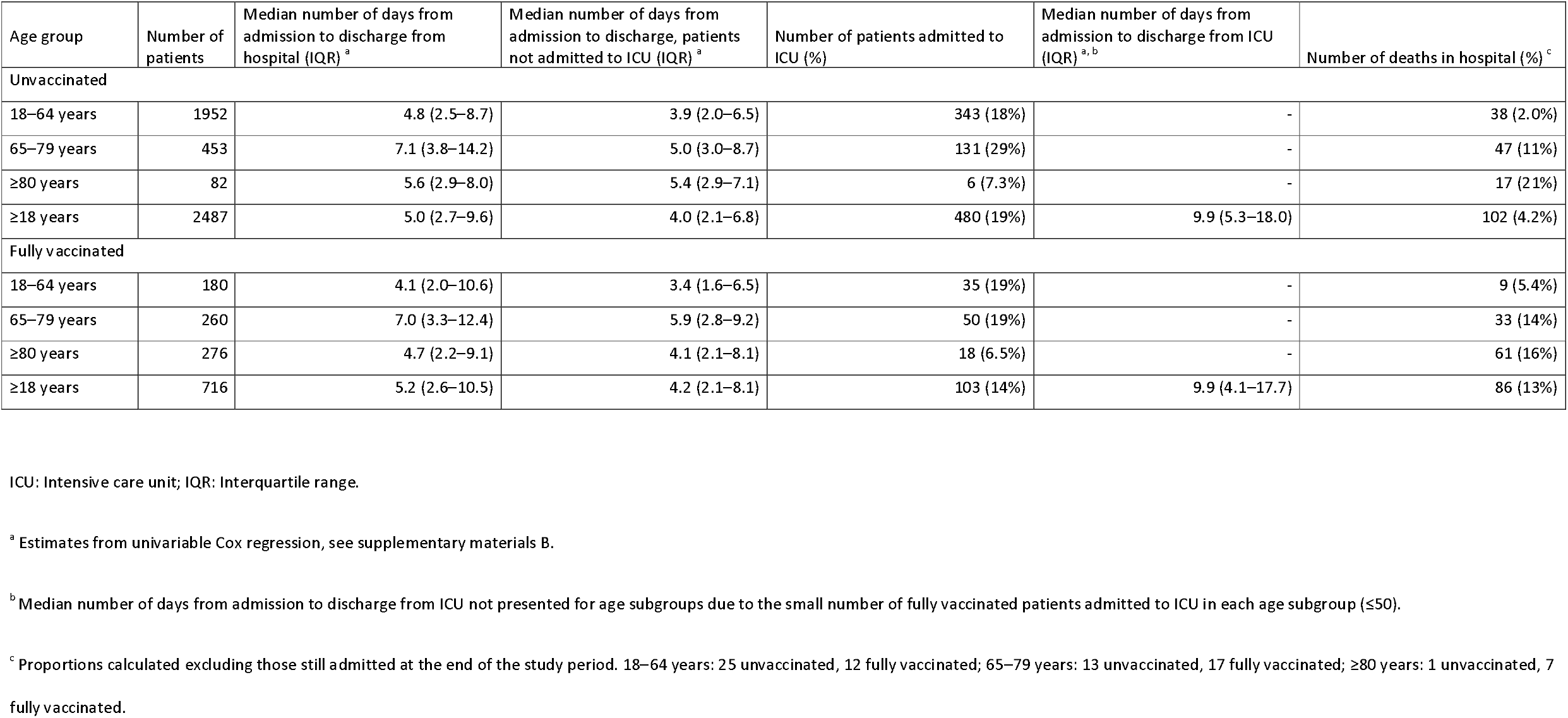
Number of patients, median number of days from admission to discharge from hospital or ICU, admissions to ICU and deaths in hospital, SARS-CoV-2 positive patients aged ≥18 years hospitalised with COVID-19 as the main cause of hospitalisation, by vaccination status and age group, Norway, 1 February – 30 November 2021

**Table 3.**
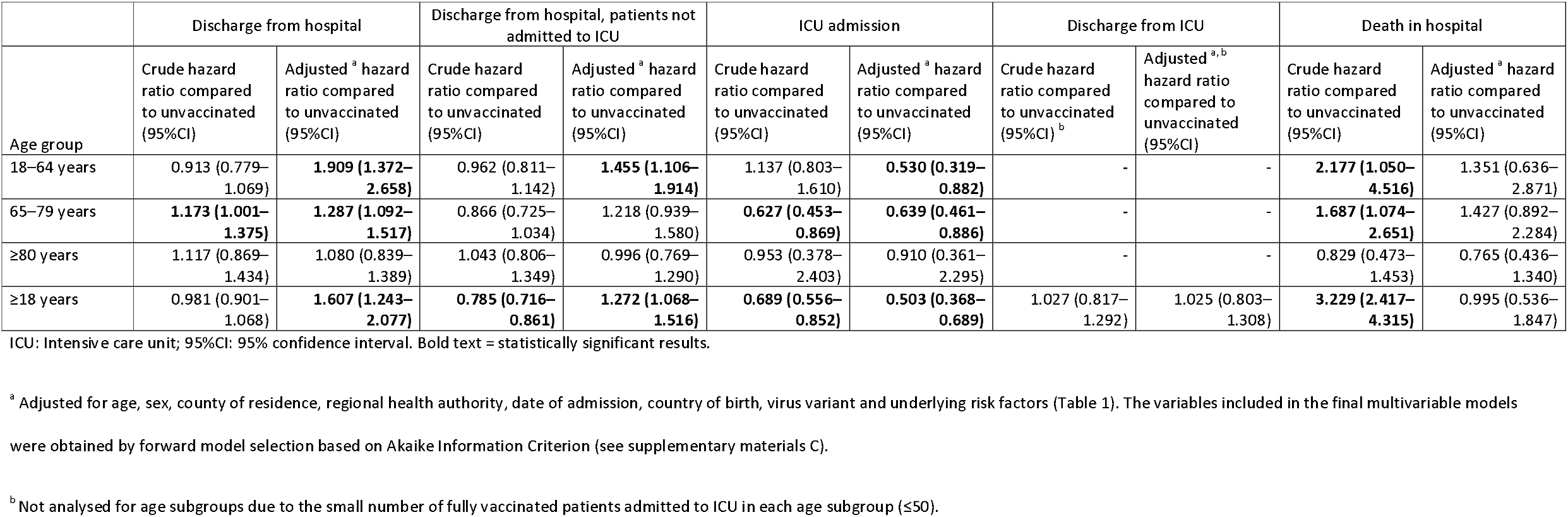
Crude and adjusted hazard ratios for discharge from hospital with and without stay in intensive care, intensive care admission, discharge from intensive care, and in-hospital death from a Cox proportional hazards model, SARS-CoV-2 positive patients aged ≥18 years hospitalised with COVID-19 as the main cause of hospitalisation, by age group, Norway, 1 February – 30 November 2021

**Figure 1.**
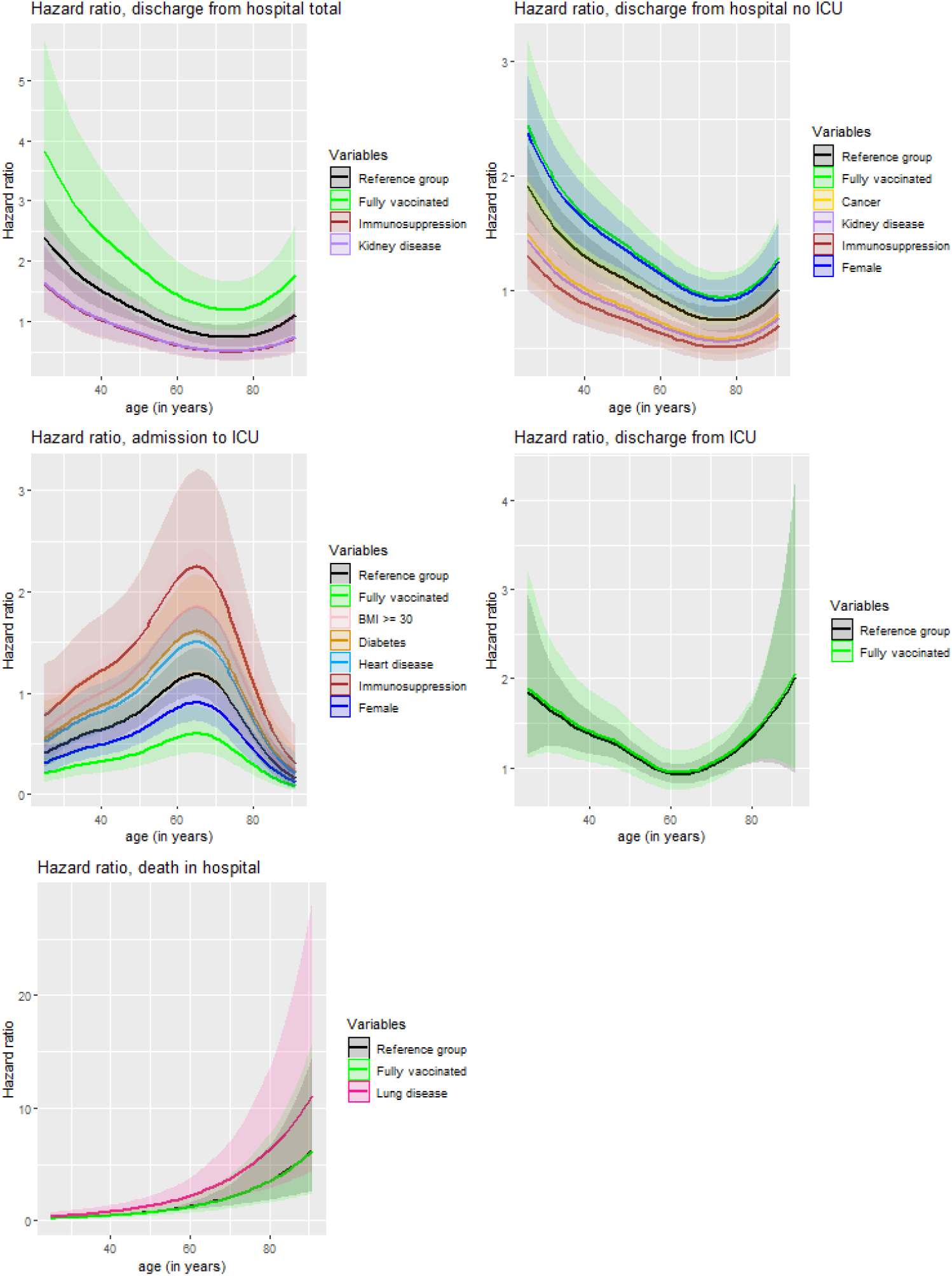
Adjusted hazard ratios for discharge from hospital with and without stay in intensive care, intensive care admission, discharge from intensive care, and in-hospital death from a Cox proportional hazards model, SARS-CoV-2 positive patients aged ≥18 years hospitalised with COVID-19 as the main cause of hospitalisation, by age, Norway, 1 February – 30 November 2021 ICU: Intensive care unit. The reference group with a hazard ratio = 1 is patients who are male, aged 56 years (median age in dataset)without underlying risk factors and unvaccinated. Hazard ratios were calculated using a Cox proportional hazards model. The variables shown in each panel are those significantly associated with each outcome in multivariable models, which were not stratified on (see supplementary materials C). Akaike Information Criterion was used for determining whether age was included linearly or with a spline.

Our multivariable models suggested that fully vaccinated patients ≥18 years had a shorter overall LoS in hospital (aHR for discharge: 1.61, 95%CI: 1.24–2.08) and shorter LoS without ICU admission (aHR: 1.28, 95%CI: 1.07–1.52) compared to unvaccinated patients. Considering exponential distribution of the survival data, an aHR for discharge of 1.61 translates into an average 38% shorter LoS (1 - 1/1.61) (18). Fully vaccinated patients also had a 50% lower risk of ICU admission (aHR: 0.50, 95%CI: 0.37– 0.69) compared to unvaccinated patients. We did not observe a difference in the LoS in ICU (aHR: 1.03, 95%CI: 0.80–1.31), or risk of in-hospital death (aHR: 1.00, 95%CI: 0.54–1.85) between vaccinated and unvaccinated patients (Figure 1, Table 3).

By age subgroup, fully vaccinated patients 18–64 years had an average 48% shorter overall LoS (aHR for discharge: 1.91, 95%CI: 1.37–2.66), 32% shorter LoS without ICU admission (aHR: 1.46, 95%CI: 1.11–1.91) and 47% lower risk of ICU admission (aHR: 0.53, 95%CI: 0.32–0.88), compared to unvaccinated individuals. Fully vaccinated patients 65–79 years had an average 22% shorter overall LoS (aHR for discharge: 1.29, 95%CI: 1.10–1.52) and 36% lower risk of ICU admission (aHR: 0.64, 95%CI: 0.46–0.89) compared to unvaccinated patients. There was no difference in the adjusted risk of in-hospital death between vaccinated and unvaccinated patients in any age subgroup. We did not observe a difference between vaccinated and unvaccinated patients ≥80 years in adjusted estimates for any outcome.

Results were generally robust in our sensitivity analyses, including when we analysed a period after which all persons in different age subgroups had been offered two vaccine doses, and when we excluded patients who had received three doses (supplementary materials A, part 2).

## Discussion

In this national register-based study, we have analysed individual-level data on 3,203 hospitalised COVID-19 patients, during a period when the majority were offered mRNA vaccines in a two-dose schedule. In line with other reports (7, 13, 14, 19), vaccinated patients were generally older and had a higher prevalence of underlying risk factors than unvaccinated patients. We find that fully vaccinated patients had a shorter LoS in hospital (both with and without ICU admission) and lower risk of ICU admission compared to unvaccinated patients. There was no difference in the LoS in ICU, or risk of in-hospital death.

Our results suggest that once hospitalised the risk of death among fully vaccinated and unvaccinated patients in Norway is similar. However, for survivors the disease trajectory is milder in fully vaccinated patients, with reduced need for hospital care and organ support. For patients not admitted to ICU, the observed reduction in LoS may have been attenuated by vaccinated patients, who may have ended up in ICU if unvaccinated, now instead spending more time in regular wards. The greatest relative reductions in LoS and risk of ICU admission were observed among patients 18– 64 years. For all outcomes, we observed no difference between vaccinated and unvaccinated patients ≥80 years. Vaccine effectiveness against hospitalisation has been reported to be lower among older age groups in Norway (20). This age group is also generally less frequently admitted to ICU, and it could be that treatment limitations confound vaccine effects in the elderly. However, the small number of unvaccinated patients ≥80 years should be considered. Our results also highlight that factors other than vaccination also continue to influence patient outcomes. A longer LoS and/or increased risk of ICU admission or death were associated with advanced age, male sex and certain risk factors such as immunosuppression, kidney disease, obesity and diabetes, as reported by others (21-24).

These findings build on previous evidence of high vaccine effectiveness against severe disease (1-5, 20) and have important implications for patient management and ongoing capacity planning in hospitals. A study including 142 patients fully vaccinated with an mRNA vaccine from 21 sites across the United States also reported a shorter LoS, lower risk of death or invasive mechanical ventilation and a lower level of clinical disease severity among vaccinated patients (13). In contrast, a study from Michigan, United States did not find lower risk of ICU admission, mechanical ventilation or death when comparing 129 fully vaccinated patients (vaccinated with Comirnaty, Spikevax or Janssen) to unvaccinated patients (14). Differences in the study cohorts, setting and design need to be considered. In this study we compare fully vaccinated and unvaccinated patients, however vaccination programmes are continuing to evolve and future analyses will be necessary to explore how parameters such as vaccine type, number of doses, time since vaccination and dose intervals affect patient outcomes between groups of vaccinated patients. While studies have suggested sustained high effectiveness of mRNA vaccines against hospitalisation at least six months following vaccination (25, 26), the duration of protection following the original two-dose schedules for mRNA vaccines and the effects of booster doses (27, 28) require ongoing research.

A strength of our study is that all data sources had national coverage. We also had a notably larger cohort of fully vaccinated patients compared to previous studies (13, 14). Also, hospitals in Norway functioned within capacity during the study period, and criteria for hospitalisation and isolation for COVID-19 patients were consistent and not related to vaccination status. Although we did not have access to treatment data, there were no major changes in treatment guidelines for COVID-19 patients in hospital or ICU in Norway during the study period. We also had minimal censoring of the study cohort, with 2.3% of patients still admitted to hospital at the end of follow-up.

Our study also has limitations. While we have controlled for several important confounders, the observational nature has the potential for residual confounding. Our fully vaccinated cohort is also predominantly representative of patients who received a homologous Comirnaty regimen. Another limitation is that some of our reported underlying risk factors do not distinguish potential differences within groups, for example whether risk factors are well-regulated or treated. Also, 40% of patients had unknown body mass index. Our model may therefore not fully adjust for certain underlying risk factors. Further, our study cohort does not include care home residents who in Norway are generally recommended to receive treatment for severe COVID-19 in their care home, not in hospital. Finally, previous natural infection has been associated with a high level of protection against SARS-CoV-2 reinfection (29, 30). While we dropped two reported reinfections, we cannot rule out that there were other previously undiagnosed SARS-CoV-2 infections in our unvaccinated cohort. If present, this may cause us to underestimate the effect of vaccination.

Our study suggests that mRNA vaccinated patients hospitalised with COVID-19 in Norway have a shorter LoS and lower risk of ICU admission than unvaccinated patients. These findings can support patient management and ongoing capacity planning in hospitals and underline the importance of vaccination programmes against COVID-19.

## Data Availability

The dataset analysed in the study contains individual-level linked data from various central health registries, national clinical registries and other national administrative registries in Norway. The researchers had access to the data through the national emergency preparedness registry for COVID-19 (Beredt C19), housed at the Norwegian Institute of Public Health (NIPH). In Beredt C19, only fully anonymised data (i.e. data that are neither directly nor potentially indirectly identifiable) are permitted to be shared publicly. Legal restrictions therefore prevent the researchers from publicly sharing the dataset used in the study that would enable others to replicate the study findings. However, external researchers are freely able to request access to linked data from the same registries from outside the structure of Beredt C19, as per normal procedure for conducting health research on registry data in Norway. Further information on Beredt C19, including contact information for the Beredt C19 project manager, and information on access to data from each individual data source, is available at https://www.fhi.no/en/id/infectious-diseases/coronavirus/emergency-preparedness-register-for-covid-19/.

## Transparency declaration

### Authors’ contributions

RW, ABK, BVS, ES, RK and EAB conceived the idea for the study. RW drafted the study protocol and coordinated the study. RK and EAB contributed directly to the acquisition of data. RW and ABK contributed to data cleaning, validation and preparation. RW and ABK led the data analysis. All co-authors contributed to the interpretation of the results. RW and ABK drafted the manuscript. All co-authors contributed to the revision of the manuscript and approved the final version for submission.

## Acknowledgements

First and foremost, we wish to thank all those who have helped establish, coordinate and report data to the national emergency preparedness registry at the Norwegian Institute of Public Health (NIPH) throughout the pandemic. We also highly acknowledge the efforts of staff at hospitals around Norway to ensure the reporting of timely and complete data to the Norwegian Intensive Care and Pandemic Registry, as well as colleagues at the register itself. We would like to specifically thank Trude Marie Lyngstad, Jostein Starrfelt, Håkon Bøås and Lamprini Veneti at the NIPH for their assistance in cleaning the data from different registries, and additionally Trude Marie Lyngstad for assistance in the production of Figure S1.

## Access to data

The dataset analysed in the study contains individual-level linked data from various central health registries, national clinical registries and other national administrative registries in Norway. The researchers had access to the data through the national emergency preparedness registry for COVID-19 (Beredt C19), housed at the Norwegian Institute of Public Health (NIPH). In Beredt C19, only fully anonymised data (i.e. data that are neither directly nor potentially indirectly identifiable) are permitted to be shared publicly. Legal restrictions therefore prevent the researchers from publicly sharing the dataset used in the study that would enable others to replicate the study findings. However, external researchers are freely able to request access to linked data from the same registries from outside the structure of Beredt C19, as per normal procedure for conducting health research on registry data in Norway. Further information on Beredt C19, including contact information for the Beredt C19 project manager, and information on access to data from each individual data source, is available at https://www.fhi.no/en/id/infectiousdiseases/coronavirus/emergency-preparedness-register-for-covid-19/.

