## supplementary materials A for "Length of hospital stay and risk of intensive care admission and in-hospital death among COVID-19 patients in Norway: a register-based cohort study comparing patients fully vaccinated with an mRNA vaccine to unvaccinated patients"

### 1. Information on the data sources

The data used in this study came from the Norwegian national emergency preparedness register, Beredt C19, which contains individual-level data from central health registries, national clinical registries and other national administrative registries. More information on Beredt C19 is available here: <https://www.fhi.no/en/id/infectious-diseases/coronavirus/emergency-preparedness-register-for-covid-19/>.

#### 1.1 Data source of hospitalised patients: the Norwegian Intensive Care and Pandemic Registry

The Norwegian Intensive Care and Pandemic Registry (NIPaR) is a national clinical registry that was expanded to include COVID-19 patients in conjunction with the COVID-19 pandemic. In the registry, all patients who have tested positive for SARS-CoV-2 and are admitted to hospital are registered. All Norwegian hospitals report to NIPaR, and reporting is mandatory. For patients who contracted SARS-CoV-2 while admitted to hospital, the time of admission is set to the date of symptom onset, or date of sampling if the patient is asymptomatic. The date of discharge for these patients is registered as when they recovered from COVID-19, even if they are still admitted to hospital for other causes. Patients who are readmitted to hospital without symptoms are only registered if they test positive for SARS-CoV-2 again and are isolated, or if a new test is not taken and the patient is isolated. Patients who are readmitted to hospital are not registered if they have tested negative for SARS-CoV-2, test positive but do not require isolation, or a new test is not taken, and the patient is not isolated. In addition, patients who are admitted to hospital for long-term complications of COVID-19 are registered if less than three months has passed since positive test. The reported main cause of hospitalisation is a clinical assessment. For patients reported with a different main cause than COVID-19, we cannot rule out that COVID-19 may have been a contributing factor for admission. In NIPaR, underlying risk factors for severe COVID-19 diagnosed before admission are registered. The following risk factors are registered; asthma, cancer, chronic lung disease, chronic neurological or neuromuscular disease, diabetes (type 1 and 2), heart disease including hypertension, immunocompromised including HIV and immunosuppressive treatment, kidney disease including kidney failure, liver disease including liver failure, pregnancy, current smoker and body mass index (calculated as weight in kilograms divided by height in centimetres squared). For cancer, only active cancer is registered, meaning cancer where the patient still receives treatment, or regular control (>1 per year). Other well-regulated or treated conditions are not distinguished from unregulated or untreated conditions, for example asthma. Full details on the registration of hospitalised patients are available here (in Norwegian): <https://helse-bergen.no/norsk-pandemiregister/registrering-i-norsk-pandemiregister-informasjon-til-ansatte>

NIPaR also includes data on patients who have tested positive for SARS-CoV-2 and are admitted to an intensive care unit (ICU). Patients are registered as ICU patients if they fulfil one of five categories:

1. Length of stay over 24 hours in intensive care
2. Require mechanical ventilation
3. Are transferred between intensive care wards
4. Persistent administration of vasoactive medication
5. Length of stay under 24 hours, but passed away during stay in intensive care

Full details on the registration of intensive care patients are available here (in Norwegian): <https://helse-bergen.no/norsk-intensivregister-nir/korona-pa-intensiv/hvordan-registrere-covid-19>

#### 1.2 Other registries included in the study

We included data on notified cases of laboratory-confirmed SARS-CoV-2 infection from the Norwegian Surveillance System for Communicable Diseases (MSIS). Data on virus variants came from the MSIS laboratory database (national laboratory database), which receives SARS-CoV-2 test results from all Norwegian microbiology laboratories. The laboratory testing for variants of SARS-CoV-2 in Norway has been described in detail elsewhere (1, 2). Data on COVID-19 vaccinations came from the Norwegian Immunisation Registry (SYSVAK). Changes overtime in national vaccination coverage for COVID-19 vaccines by age group and number of doses are presented in Figure S1. Data on persons with a national identity number was drawn from the national population registry. The national identity number was essential to link data from all registries used in the analysis.

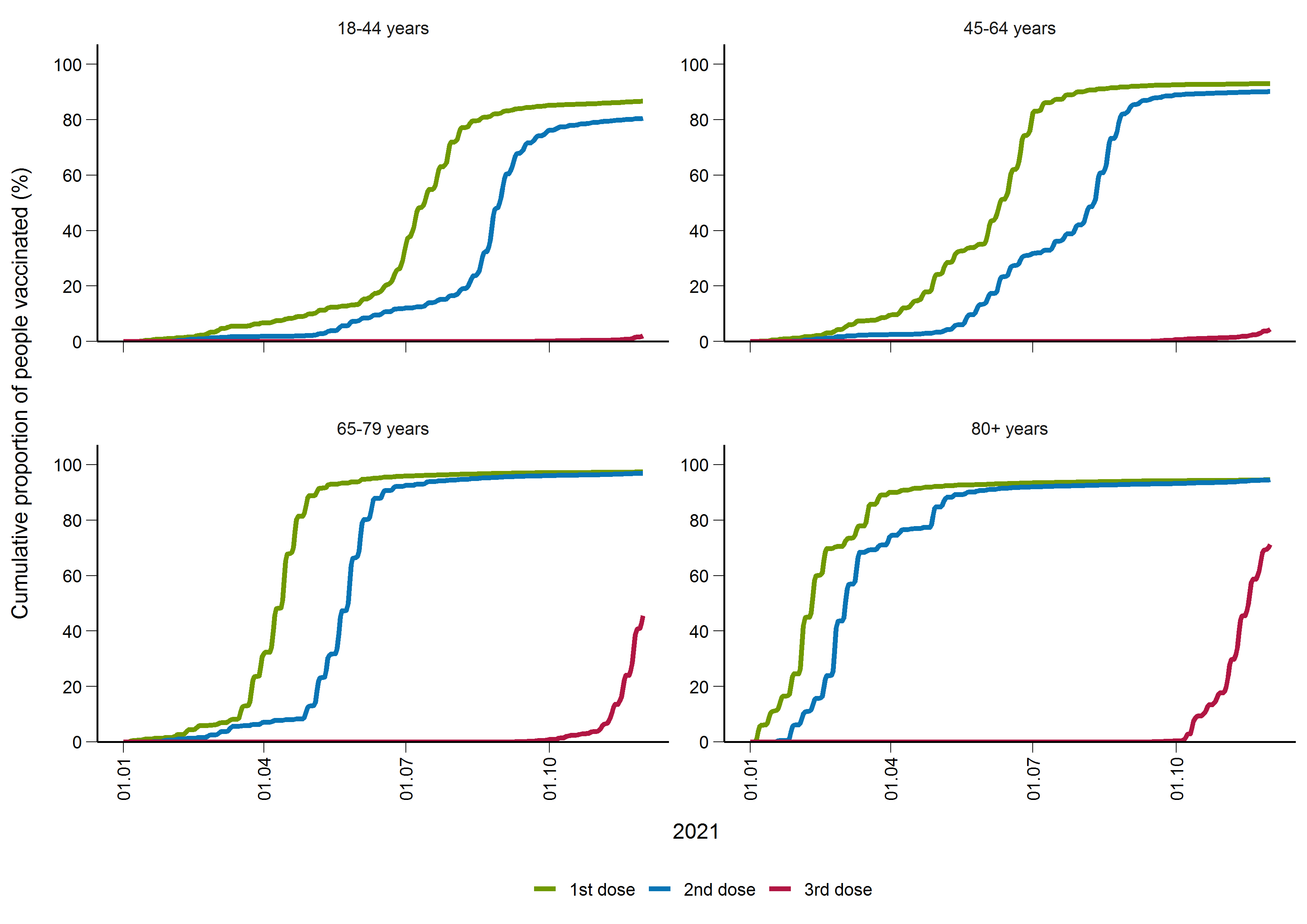

*Figure S1. National vaccination coverage of COVID-19 vaccines in Norway by age group, number of doses and date, 1 January – 30 November 2021.*

The age group 18–64 years is split into 18–44 and 45–64 years in Figure S1, as vaccination of the general population started later in those 18–44 in Norway (see (3)).

### 2. Sensitivity analyses

In addition to the main analysis, we conducted sensitivity analyses by changing the definition of our study population, period of analysis (to take into account progression of the vaccination programme in different age groups (3)) or our outcome definitions to further explore if our main results were robust. The adjusted estimates from our sensitivity analyses are presented in Table S1. The methodology used was the same as the one in the main analysis in each age group (see manuscript and supplementary material C). We did not conduct sensitivity analyses on the outcome length of stay in ICU.

Results were generally robust in all our sensitivity analyses, including when we adjusted the period of analysis to a time after which all persons in different age groups had been offered two vaccine doses.

Among the age group 18–64 years we observed a lower risk of death among fully vaccinated patients in several different sensitivity analyses where we adjusted our study population. For example, when excluding patients who had received three vaccine doses and fully vaccinated patients with >180 days between date of last dose and positive test, or when including patients who had received one dose <21 days before positive test as unvaccinated, and patients who had received one dose ≥21 days before positive test as vaccinated. This reflects that it is likely only 18–64-year-olds in high-risk groups who would have received a third dose or received their last dose more than 180 days ago in our study cohort, while 18–64-year-olds who had only received one dose are less likely to belong to high-risk groups and vaccination with a first dose <21 days before positive test may not be enough time to develop sufficient protection again severe disease.

When including all SARS-CoV-2 positive patients regardless of main cause of hospitalisation, fully vaccinated patients 18–64 years and ≥80 years had a lower risk of in-hospital death. For unvaccinated patients with another main cause of hospitalisation, COVID-19 may have been a more significant contributing factor for admission, while frail elderly patients with multiple comorbidities may be more likely to be unvaccinated, as potential side effects can outweigh the protection offered by vaccination in this group.

Among 18–64-year-olds we also observed no decrease in the length of stay for patients not admitted to ICU when excluding patients with a length of stay <24 hours, contrary to our main analysis. This highlights the proportionally greater number of short non-ICU stays among fully vaccinated patients in this age group, although the 95%CI was on the cusp of statistical significance.

Finally, in patients ≥18 years when excluding those who had received three vaccine doses and fully vaccinated patients with >180 days between date of last dose and positive test we observed no difference in the length of stay in hospital overall between fully vaccinated and unvaccinated patients. This is likely related to how this sensitivity analysis affects the study population ≥18 years. By excluding those who had received three vaccine doses and fully vaccinated patients with >180 days between date of last dose and positive test, we are removing the majority of vaccinated patients aged ≥80 years, who generally have a shorter length of stay than patients aged 55–79 years, the age group which accounts for the majority of vaccinated patients <80 years. In the same sensitivity analysis for the age subgroups 18–64 years, 65–79 years and ≥80 years we did observe shorter lengths of stay overall, as well as a shorter length of stay without ICU and lower risk of ICU admission among patients ≥18 years. For those aged 18–64 years and 65–79 years this reflects the results of the main analysis. For ≥80 years a shorter length of stay when excluding such patients may indicate waning immunity among those vaccinated more than 6 months ago.

In our study cohort, 583 patients were admitted to ICU and 188 died. Of those 188, 106 had been admitted to ICU, while 82 died without admission to ICU. In addition to the sensitivity analyses presented in Table S1, we also ran a model where the outcome was admission to ICU or death in hospital (n=665). Among patients aged 18–64 years (aHR 0.577, 95%CI 0.349–0.953) and 65–79 years (aHR 0.722, 95%CI 0.536–0.972) results reflected those for admission to ICU while for patients aged ≥80 years (aHR 0.730, 95%CI 0.443–1.203) results more closely reflected those for death in hospital.

*Table S1. Sensitivity analyses with adjusted hazard ratios for discharge from hospital with and without stay in intensive care, intensive care admission and in-hospital death from a Cox proportional hazards model, by vaccination status and age group, Norway, 1 February – 30 November 2021.*

| Age group | Vaccination status | Discharge from hospital | | Discharge from hospital, patients not admitted to ICU | | Admission to ICU | | Death in hospital | |
| --- | --- | --- | --- | --- | --- | --- | --- | --- | --- |
|  |  | Number of patients | Adjusted hazard ratio (95%CI) | Number of patients | Adjusted hazard ratio (95%CI) | Yes | Adjusted hazard ratio (95%CI) | Yes | Adjusted hazard ratio (95%CI) |
| Main analysis | | | | | | | | | |
| 18–64 years | Unvaccinated | 1952 | Ref | 1609 | Ref | 343 | Ref | 38 | Ref |
|  | Fully vaccinated | 180 | **1.909 (1.372–2.658)** | 145 | **1.455 (1.106–1.914)** | 35 | **0.530 (0.319–0.882)** | 9 | 1.351 (0.636–2.871) |
| 65–79 years | Unvaccinated | 453 | Ref | 322 | Ref | 131 | Ref | 47 | Ref |
|  | Fully vaccinated | 260 | **1.287 (1.092–1.517)** | 210 | 1.218 (0.939–1.580) | 50 | **0.639 (0.461–0.886)** | 33 | 1.427 (0.892–2.284) |
| ≥80 years | Unvaccinated | 82 | Ref | 76 | Ref | 6 | Ref | 17 | Ref |
|  | Fully vaccinated | 276 | 1.080 (0.839–1.389) | 258 | 0.996 (0.769–1.290) | 18 | 0.910 (0.361–2.295) | 44 | 0.765 (0.436–1.340) |
| ≥18 years | Unvaccinated | 2487 | Ref | 2007 | Ref | 480 | Ref | 102 | Ref |
|  | Fully vaccinated | 716 | **1.607 (1.243–2.077)** | 613 | **1.272 (1.068–1.516)** | 103 | **0.503 (0.368–0.689)** | 86 | 0.995 (0.536–1.847) |
| Sensitivity analyses | | | | | | | | | |
| Excluding patients who had received three vaccine doses | | | | | | | | | |
| 18–64 years | Unvaccinated | 1952 | Ref | 1609 | Ref | 343 | Ref | 38 | Ref |
|  | Fully vaccinated | 167 | **1.827 (1.307–2.555)** | 137 | **1.395 (1.057–1.841)** | 30 | **0.453 (0.265–0.773)** | 9 | **0.516 (0.307–0.870)** |
| 65–79 years | Unvaccinated | 453 | Ref | 322 | Ref | 131 | Ref | 47 | Ref |
|  | Fully vaccinated | 242 | **1.305 (1.104–1.541)** | 198 | 1.228 (0.948–1.592) | 44 | **0.600 (0.426–0.845)** | 26 | 1.178 (0.712–1.948) |
| ≥80 years | Unvaccinated | 82 | Ref | 76 | Ref | 6 | Ref | 17 | Ref |
|  | Fully vaccinated | 260 | 1.088 (0.844–1.402) | 244 | 1.000 (0.770–1.297) | 16 | 0.862 (0.337–2.205) | 40 | 0.751 (0.425–1.327) |
| ≥18 years | Unvaccinated | 2487 | Ref | 2007 | Ref | 480 | Ref | 102 | Ref |
|  | Fully vaccinated | 669 | **1.611 (1.240–2.093)** | 579 | **1.259 (1.055–1.502)** | 90 | **0.473 (0.342–0.653)** | 75 | 0.955 (0.504–1.809) |
| Excluding patients who had received three vaccine doses and fully vaccinated patients with >180 days between date of last dose and positive test | | | | | | | | | |
| 18–64 years | Unvaccinated | 1952 | Ref | 1609 | Ref | 343 | Ref | 38 | Ref |
|  | Fully vaccinated | 136 | **1.816 (1.257–2.622)** | 111 | **1.406 (1.043–1.896)** | 25 | **0.489 (0.277–0.862)** | 9 | **0.550 (0.317–0.954)** |
| 65–79 years | Unvaccinated | 453 | Ref | 322 | Ref | 131 | Ref | 47 | Ref |
|  | Fully vaccinated | 158 | **1.345 (1.111–1.628)** | 130 | 1.197 (0.915–1.566) | 28 | **0.571 (0.379–0.859)** | 17 | 1.218 (0.686–2.163) |
| ≥80 years | Unvaccinated | 82 | Ref | 76 | Ref | 6 | Ref | 17 | Ref |
|  | Fully vaccinated | 49 | **1.727 (1.196–2.493)** | 47 | **1.790 (1.225–2.616)** | 2 | 0.689 (0.138–3.454) | 3 | 0.373 (0.102–1.363) |
| ≥18 years | Unvaccinated | 2487 | Ref | 2007 | Ref | 480 | Ref | 102 | Ref |
|  | Fully vaccinated | 343 | 1.286 (0.914–1.811) | 288 | **1.370 (1.110–1.690)** | 55 | **0.477 (0.331–0.689)** | 29 | 0.676 (0.289–1.579) |
| Including 156 additional patients who had received one dose <21 days before positive test as unvaccinated, and 94 additional patients who had received one dose ≥21 days before positive test as vaccinated | | | | | | | | | |
| 18–64 years | Unvaccinated | 2040 | Ref | 1679 | Ref | 361 | Ref | 38 | Ref |
|  | Partially or fully vaccinated | 237 | **2.013 (1.518–2.670)** | 198 | **1.596 (1.262–2.018)** | 39 | **0.487 (0.305–0.777)** | 10 | **0.536 (0.339–0.847)** |
| 65–79 years | Unvaccinated | 508 | Ref | 362 | Ref | 146 | Ref | 55 | Ref |
|  | Partially or fully vaccinated | 288 | **1.228 (1.052–1.435)** | 234 | 1.051 (0.838–1.319) | 54 | **0.611 (0.447–0.835)** | 38 | 1.289 (0.835–1.990) |
| ≥80 years | Unvaccinated | 95 | Ref | 88 | Ref | 7 | Ref | 21 | Ref |
|  | Partially or fully vaccinated | 285 | 1.107 (0.872–1.405) | 266 | 1.042 (0.815–1.332) | 19 | 0.979 (0.411–2.332) | 47 | 0.816 (0.486–1.372) |
| ≥18 years | Unvaccinated | 2643 | Ref | 2129 | Ref | 514 | Ref | 114 | Ref |
|  | Partially or fully vaccinated | 810 | **1.549 (1.224–1.961)** | 698 | **1.281 (1.093–1.503)** | 112 | **0.489 (0.365–0.655)** | 95 | 0.930 (0.540–1.602) |
| Study time period June to November–instead of February to November (two vaccine doses had been offered to all age groups ≥65 years in Norway by the end of May) | | | | | | | | | |
| 65–79 years | Unvaccinated | 117 | Ref | 81 | Ref | 36 | Ref | 16 | Ref |
|  | Fully vaccinated | 256 | **1.531 (1.203–1.950)** | 206 | 1.249 (0.949–1.645) | 50 | **0.630 (0.409–0.968)** | 32 | 1.104 (0.601–2.029) |
| Study time period September to November–instead of February to November (two vaccine doses had been offered to all age groups ≥18 years in Norway in early September) | | | | | | | | | |
| 18–64 years | Unvaccinated | 400 | Ref | 314 | Ref | 86 | Ref | 10 | Ref |
|  | Fully vaccinated | 160 | **2.185 (1.489–3.207)** | 128 | **1.421 (1.040–1.941)** | 32 | **0.512 (0.275–0.952)** | 6 | 0.551 (0.300–1.012) |
| ≥18 years | Unvaccinated | 538 | Ref | 415 | Ref | 123 | Ref | 30 | Ref |
|  | Fully vaccinated | 639 | **1.716 (1.265–2.329)** | 544 | **1.410 (1.142–1.741)** | 95 | **0.488 (0.338–0.704)** | 76 | 0.977 (0.499–1.914) |
| Including all patients hospitalised after a positive SARS-CoV-2 test, regardless of main cause of hospitalisation | | | | | | | | | |
| 18–64 years | Unvaccinated | 2336 | Ref | 1969 | Ref | 367 | Ref | 44 | Ref |
|  | Fully vaccinated | 242 | **1.616 (1.259–2.073)** | 207 | **1.290 (1.036–1.606)** | 35 | **0.468 (0.288–0.760)** | 12 | **0.505 (0.315–0.810)** |
| 65–79 years | Unvaccinated | 522 | Ref | 384 | Ref | 138 | Ref | 56 | Ref |
|  | Fully vaccinated | 352 | **1.286 (1.113–1.487)** | 299 | 1.199 (0.951–1.513) | 53 | **0.540 (0.393–0.742)** | 38 | 1.186 (0.770–1.828) |
| ≥80 years | Unvaccinated | 119 | Ref | 106 | Ref | 13 | Ref | 34 | Ref |
|  | Fully vaccinated | 376 | 1.144 (0.927–1.411) | 352 | 1.041 (0.836–1.296) | 24 | 0.651 (0.331–1.281) | 60 | **0.605 (0.396–0.925)** |
| ≥18 years | Unvaccinated | 2977 | Ref | 2459 | Ref | 518 | Ref | 134 | Ref |
|  | Fully vaccinated | 970 | **1.459 (1.216–1.750)** | 858 | **1.209 (1.056–1.385)** | 112 | **0.463 (0.345–0.622)** | 110 | 0.959 (0.587–1.565) |
| Excluding patients who had a length of stay <24 hours | | | | | | | | | |
| 18–64 years | Unvaccinated | 1770 | Ref | 1427 | Ref | 343 | Ref | 38 | Ref |
|  | Fully vaccinated | 156 | **1.694 (1.179–2.435)** | 121 | 1.305 (0.962–1.771) | 35 | **0.529 (0.315–0.890)** | 9 | **0.596 (0.359–0.990)** |
| 65–79 years | Unvaccinated | 437 | Ref | 306 | Ref | 131 | Ref | 47 | Ref |
|  | Fully vaccinated | 251 | **1.308 (1.106–1.547)** | 201 | 1.203 (0.923–1.567) | 50 | **0.640 (0.462–0.887)** | 32 | 1.374 (0.855–2.207) |
| ≥80 years | Unvaccinated | 78 | Ref | 73 | Ref | 5 | Ref | 14 | Ref |
|  | Fully vaccinated | 251 | 1.046 (0.807–1.356) | 233 | 0.946 (0.724–1.235) | 18 | 1.118 (0.415–3.013) | 43 | 0.908 (0.496–1.663) |
| ≥18 years | Unvaccinated | 2285 | Ref | 1806 | Ref | 479 | Ref | 99 | Ref |
|  | Fully vaccinated | 658 | **1.496 (1.140–1.963)** | 555 | 1.193 (0.991–1.435) | 103 | **0.523 (0.382–0.718)** | 84 | 1.102 (0.585–2.076) |
| Excluding all time between hospital stays in calculating total length of stay (in the main analysis we included time between stays as part of the patient’s length of stay, if the time between two consecutive stays was less than 24 hours, to allow for transfers between hospitals). | | | | | | | | | |
| 18–64 years | Unvaccinated | 1952 | Ref | 1609 | Ref | 343 | Ref | 38 | Ref |
|  | Fully vaccinated | 180 | **1.910 (1.372–2.658)** | 145 | **1.455 (1.106–1.914)** | 35 | **0.512 (0.306–0.855)** | 9 | 0.972 (0.688–1.373) |
| 65–79 years | Unvaccinated | 453 | Ref | 322 | Ref | 131 | Ref | 47 | Ref |
|  | Fully vaccinated | 260 | **1.288 (1.093–1.518)** | 210 | 1.222 (0.942–1.586) | 50 | **0.639 (0.461–0.886)** | 33 | 1.427 (0.892–2.283) |
| ≥80 years | Unvaccinated | 82 | Ref | 76 | Ref | 6 | Ref | 17 | Ref |
|  | Fully vaccinated | 276 | 1.079 (0.839–1.387) | 258 | 0.996 (0.769–1.291) | 18 | 0.910 (0.361–2.295) | 44 | 0.765 (0.436–1.341) |
| ≥18 years | Unvaccinated | 2487 | Ref | 2007 | Ref | 480 | Ref | 102 | Ref |
|  | Fully vaccinated | 716 | **1.609 (1.245–2.080)** | 613 | **1.273 (1.068–1.517)** | 103 | **0.503 (0.368–0.689)** | 86 | 0.996 (0.537–1.850) |
| Including deaths up to 7 days post-discharge in analysis of risk for death, instead of only death in hospital as in the main analysis. Only patients discharged by 6 December are included to ensure 7 days of follow-up. | | | | | | | | | |
| ≥80 years | Unvaccinated | - | - | - | - | - | - | 23 | Ref |
|  | Fully vaccinated | - | - | - | - | - | - | 56 | 0.723 (0.444–1.176) |
| ≥18 years | Unvaccinated | - | - | - | - | - | - | 108 | Ref |
|  | Fully vaccinated | - | - | - | - | - | - | 99 | 0.924 (0.505–1.692) |

ICU: Intensive care unit; 95%CI: 95% confidence interval. Bold text = statistically significant results. Red text = associations that differ in statistical significance from the main analysis.

### 3. Distribution of vaccine type in the study cohort

*Table S2: Distribution of vaccine type among fully vaccinated SARS-CoV-2 patients in the study cohort*

| Vaccine type | One dose + previous COVID-19 infection | Two doses | Three doses |
| --- | --- | --- | --- |
| Comirnaty | 3 | 618 | 37 |
| Spikevax | 0 | 32 | 1 |
| Comirnaty + Spikevax | - | 6 | - |
| Vaxzevria + Comirnaty | - | 10 | - |
| Comirnaty + Comirnaty + Spikevax | - | - | 9 |
| Total | 3 | 666 | 47 |

*Table S3: Distribution of time in weeks between dose one and two among SARS-CoV-2 patients vaccinated with at least two doses of a COVID-19 vaccine in the study cohort*

|  | Vaccine type | | | |
| --- | --- | --- | --- | --- |
| Number of weeks between doses | Dose one: Comirnaty Dose two: Comirnaty | Dose one: Spikevax Dose two: Spikevax | Dose one: Comirnaty Dose two: Spikevax | Vaxzevria + Comirnaty |
| <3 ^a^ | 8 | 0 | 0 | 0 |
| 3 | 304 | 13 | 0 | 0 |
| 4 | 17 | 2 | 1 | 0 |
| 5 | 29 | 16 | 0 | 0 |
| 6 | 280 | 2 | 2 | 1 |
| 7 | 10 | 0 | 0 | 0 |
| 8 | 6 | 0 | 2 | 0 |
| 9 | 2 | 0 | 1 | 1 |
| 10 | 3 | 0 | 0 | 2 |
| 11 | 1 | 0 | 0 | 1 |
| 12 | 0 | 0 | 0 | 5 |
| 13 | 0 | 0 | 0 | 0 |
| ≥14 | 4 ^b^ | 0 | 0 | 0 |
| Total | 664 | 33 | 6 | 10 |

^a^ Dose interval of 19 or 20 days, as per the absolute minimum interval between doses for Comirnaty (4).

^b^ Two 14 weeks, one 31 weeks, one 39 weeks.

*Table S4: Distribution of time in weeks between dose two and three among SARS-CoV-2 patients vaccinated with three doses of a COVID-19 vaccine in the study cohort*

|  | Vaccine type | | |
| --- | --- | --- | --- |
| Number of weeks between doses | Dose two: Comirnaty Dose three: Comirnaty | Dose two: Spikevax Dose three: Spikevax | Dose two: Comirnaty Dose three: Spikevax |
| 6-10 | 2 | 0 | 0 |
| 11-15 | 0 | 0 | 1 |
| 16-20 | 12 | 0 | 3 |
| 21-25 | 4 | 0 | 3 |
| 26-30 | 7 | 1 | 2 |
| 31-35 | 8 | 0 | 0 |
| 36-40 | 4 | 0 | 0 |
| Total | 37 | 1 | 9 |
